# Rheumatic Heart Disease Detection in Asymptomatic Schoolchildren using ECG and PCG

**DOI:** 10.64898/2026.05.12.26352939

**Authors:** Amsalu Tomas Chuma, Chunzhuo Wang, Jens-Uwe Voigt, Desalew Mekonnen Kassie, Melkamu Hunegnaw Asmare, Bart Vanrumste

## Abstract

**Background:** Rheumatic heart disease (RHD) remains a major public health concern across low- and middle-income countries in the Global South. Early detection through community-based screening of asymptomatic individuals has been identified as a critical strategy for reducing the disease burden. Despite this, the absence of accessible, automated population screening tools continues to impede implementation at scale. This study investigates the screening potential of integrating electrocardiography (ECG) and phonocardiography (PCG) for the early detection of RHD in asymptomatic schoolchildren.

**Methods:** The dataset was obtained as part of an ambulatory screening initiative conducted across multiple school sites in rural areas of Ethiopia. It comprised ECG and PCG recordings from 611 asymptomatic schoolchildren aged 10 to 20 years. A comprehensive set of time–frequency, visibility graph and non-linear features were extracted from both signal modalities. These features were subsequently evaluated using machine learning models to assess their utility in the automated screening of early RHD.

**Results:** The best model achieved an average 10-folds cross-validation scores on sensitivity, positive-predictive-value and F1-score of 59.6%, 63.6% and 60.8%, respectively for multimodal ECG and PCG signals. Whereas separate evaluation of ECG showed an F1-score of 61.1% and PCG achieved 23.5%. Key features included the T-wave, the area under the QRS complex, and entropy measures derived from beat visibility graphs in the ECG. In addition, visibility graph features from multi-band S1 and S2 heart sound segments, along with MFCC coefficients from the PCG, were also relevant. However, PCG alone performed poorly and did not show improved results over the ECG features.

**Conclusion:** Although auscultation is key clinical diagnosis tool in symptomatic RHD, combined PCG with ECG features does not enhance asymptomatic RHD detection using the ECG modality alone.

## 1. Introduction

### 1.1 Rheumatic Heart Disease and its Diagnostic Challenges

Subclinical Rheumatic Heart Disease (RHD) remains a significant public health concern in low- and middle-income countries (LMICs) mainly due to the lack of medical resources, trained personnel and socio-economic barriers [1]. The RHD associated health burden in LIMCs shows no major decline in the past decades [2, 3]. Hospital-based studies reported that RHD accounts for 6.6 to 34.0% of hospital admissions related to cardiovascular diseases [4–6]. RHD is the chronic manifestation of acute rheumatic fever (ARF), often resulting from an autoimmune response to Group A Streptococcus infection. Schoolchildren and young adults in at-risk areas are more prone to recurrent ARF and consequently develop RHD. The disease leads to valvular deformities, predominantly affecting the mitral valve, followed by aortic involvement [7]. Overtime, these structural changes generate distinctive murmurs in auscultatory and abnormalities in electrocardiograms (ECG) [1].

The World Heart Federation (WHF) criteria [7, 8] established echocardiography as the definitive diagnostic tool for RHD diagnosis. Doppler modalities further used to quantify the severity of regurgitant or stenotic lesions [7]. In contrast, auscultation-based diagnosis reported poor performance compared to echocardiography in screening for RHD among children with about 38.2% sensitivity and 75.1% specificity [9–13]. In addition, ECG analysis in symptomatic RHD patients has also been reported to exhibit rhythm disturbances and prolonged non-specific intervals such as PR, QTc and QRS interval [14–18]. Particularly, in acute inflammatory phase of the disease, about 70% of the patients show rheumatic endocarditis which alters the T-wave and TpTe parameters [14, 15].

Although echocardiography significantly outperforms auscultation [9, 11, 12], it is costly and infrastructure-intensive for majority of the disease endemic countries. Moreover, access to high-quality echocardiography equipment and trained sonographers is often limited, making mass screening for RHD to be impractical [13]. Even with the advent of handheld echo devices, interpretation remains a bottleneck without adequate cardiology support.

### 1.2 Heart Murmurs in RHD

The RHD caused regurgitant valves create turbulent blood flow, which can be heard as heart murmurs in phonocardiogram (PCG). Example PCG and ECG recordings from confirmed RHD subjects are shown in Figures 1(B) and 1(D), respectively, with comparative signals from a healthy subject in Figures 1(A) and 1(C). The mitral regurgitation (MR) murmurs are typically holosystolic with high-frequency components peaking around 200–400 Hz beginning at S1 and continues up to S2 [19, 20]. The mitral stenosis (MS) murmurs are low frequency diastolic rumbles around 20–200Hz begins soon after S2 sound following the opening snap and lasts through mid to late diastole. Aortic Regurgitation (AR) is an early diastolic decrescendo murmur within frequency range of 300–600Hz [19, 20].

**Figure 1.**
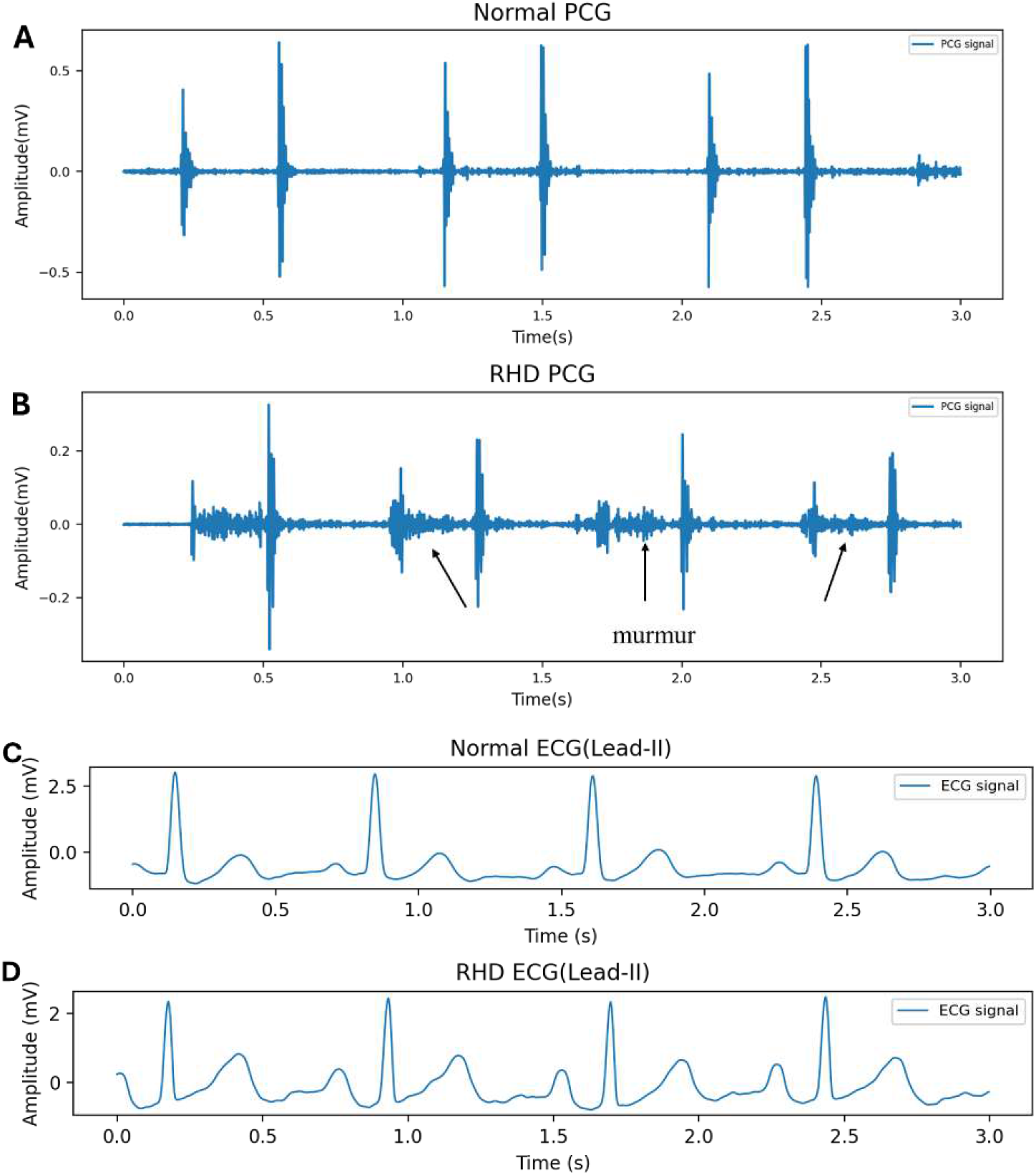
Example PCG and ECG recordings. (A) PCG recording from Normal participant, (B) PCG recording from RHD participant. And in (C) an ECG recording from confirmed RHD participant

Such pathologic murmurs are distinct from innocent murmurs [7], which are usually mid-systolic, soft, and limited in spectral bandwidth. In schoolchildren aged between 5 to 12 years, however, pathologic murmurs may be softer due to lower cardiac output and thinner thoracic walls, while adolescents between 13 to 19 years of age show more pronounced patterns due to increased cardiac mass and systemic pressure [21, 22]. In addition, PCG depends on hemodynamically significant valvular turbulence, which is better revealed during the later manifestation, not as an inflammatory marker at early stage [21]. These features attribute to low sensitivity of auscultation for screening RHD in resource limited settings [11, 23]. Thus, accurate differentiation requires analysing time-aligned spectral and temporal PCG patterns that can be aided by machine learning (ML) for improved detection.

Many studies have investigated automated heart sound classification using ML and deep learning techniques, often benchmarking on annotated phonocardiogram datasets such as PhysioNet CinC 2016 and 2022 [24, 25]. Early works focused on time-domain features like amplitude, timing intervals, zero-crossing rate, energy, entropy, kurtosis, and skewness to distinguish murmurs using ML classifiers [26]. Frequency-domain features, including power spectral density (PSD), discrete wavelet transform (DWT), continuous wavelet transform (CWT), Mel-frequency cepstral coefficients (MFCCs), and spectral centroid, further improved performance [27–31]. Additionally, nonlinear and chaotic dynamics analyses revealed that healthy hearts exhibit more complex chaotic behaviour compared to abnormal ones [32, 33]. Particularly in advanced symptomatic RHD, these features showed a sensitivity and specificity of 98% and 95%, respectively [33].

A lightweight Convolutional Neural Networks (CNN) were also used to classify abnormal heart murmurs (AS, MR, MS, and mitral valve prolapse) from normal, achieving an accuracy of 98.6% in a balanced dataset combined from CinC 2016 and PASCAL datasets [34]. The top-performed methods in these challenges combined hand-engineered features with deep learning architectures such as CNN or Long Short-Term Memory networks (LSTM), achieving notable gains [35–39]. These hybrid models fused domain-informed features with learned representations, enhancing robustness and generalization especially under class imbalance and noisy conditions.

### 1.3 Electrocardiographic patterns in RHD

On the other hand, patients with subclinical RHD may have no clinically apparent functional cardiac abnormalities [40], and ECG changes may therefore be absent or subtle, leading to non-specific ECG patterns. While the PR interval and P-wave duration changes were reported in early-stage RHD [14, amsalu3, amsalu4], the prolongation of PR and QTc intervals [14], and T-wave changes were reported [15, 16, 17]. Wearable ECG devices, such as KardiaMobile, provide portable and convenient cardiac monitoring by capturing high-quality single-lead ECG signals outside clinical environments. Automated detection of various arrhythmia from single-lead ECGs of 53,549 subjects and 12-lead ECGs from more than 1,558,848 recordings using deep learning was reported at human-level [41,42].

Studies have demonstrated that deep neural networks can effectively identify arrhythmias due to functional heart diseases by analyzing raw ECG signals, achieved a human-level performance [41, 43]. In contrast, the use of ECG for detecting structural heart diseases such as RHD, and its associated manifestations is less established. Deep learning-based approaches indicated the extraction of subtle patterns from ECGs indicative of left ventricular dysfunction [44], severe aortic regurgitation [45], and mitral regurgitation [46]. In particular, the work of [47] reported a sensitivity of 90% and specificity of 53.3% for MR detection. A relatively larger study [48] used a multi-input model comprised a two-dimensional convolutional neural network (2D-CNN) using raw ECG data and a fully connected deep neural network (FC-DNN) using ECG features for severe aortic regurgitation. The authors reported sensitivity of 53.5%, and specificity of 82.8% on a dataset that consisted of 29,859 paired ECG and echocardiography. Similarly, the work of [45] proposed an ECG-based screening for valvular heart disease using ValveNet model which utilized a 1D CNN architecture to analyze 12-lead ECGs from 77,163 patients, achieved an area under the receiver operating characteristic curve (AUROC) of 0.88 for AS, 0.77 for AR, and 0.83 for MR.

While these deep learning models largely operate on raw signals, hybrid approaches improve the performances by combining hand-engineered time-frequency features such as wavelets, power spectral density (PSD), and entropy [49, 50]. Nevertheless, in RHD, morphological changes typically emerge only in advanced stages [7, 18], whereas rhythmic irregularities and prolonged temporal intervals are reported during the early phase of the disease [51]. Therefore, integrating features derived from both multimodal ECG and PCG signals could offer a complementary approach for capturing early functional disruptions and the murmurs.

In this work, we leverage time-frequency, graph-based and non-linear features extracted from randomly selected windowed frames of both PCG and ECG signals, as well as from four fundamental heart sounds (FHS) including S1, S2, systolic and diastolic segments using the algorithm explained in [52]. Different ML models were evaluated on these set of features to classify between healthy and individuals with RHD (PwRHD) among asymptomatic at-risk schoolchildren. The advantage of combining the PCG with the ECG features on the model performance was assessed. The contributions of this paper are four-fold: First, we propose a novel approach for early detection of RHD using multimodal, low-cost ECG and PCG sensors. Second, we introduce a comprehensive set of features extracted from both modalities. Additionally, we assess the individual contributions of each modality and demonstrate the effectiveness of their combination for RHD screening in endemic regions. Finally, we present an ECG and PCG dataset recorded from asymptomatic schoolchildren screened for RHD. The ECG and PCG were not measured synchronously but rather subsequently.

## 2. Materials and Methods

### 2.1 RHDECG dataset

The ECG dataset was explained in our earlier work [51]. A total of 611 30-seconds duration ECGs recorded using a commercial CE marked device, KardiaMobile™ 6L (AliveCore Inc., Mountain View, California, USA) (KM) with a sampling frequency of 300Hz. The recordings were from asymptomatic schoolchildren during the RHD screening campaign, consisting of 564 healthy controls and 47 participants with confirmed RHD. The “kardia app” from AliveCor Inc. was used to connect the sensor and a smartphone via Bluetooth for recording, and consequently saved to computer for further analysis. Six leads, Leads (I, II, III, aVR, aVF, aVL), were extracted for analysis (see Figure 1(b)).

### 2.2 RHDPCG dataset

The PCG dataset was recorded using ThinkLabs One™ stethoscope (ThinkLabs, Colorado, USA), from the same study cohort as the RHDECG dataset. The device was connected to a computer for storing the files using Audacity software (v3.7.2). During acquisition of heart sounds, the frequency filter of the stethoscope was set to wideband between [20 – 20,000] Hz. As described in [51], the recordings were from four auscultatory positions, namely aortic, pulmonic, tricuspid and mitral areas. The sampling rate was 14.1kHz. The mitral and tricuspid area recordings typically exhibit higher and more pronounced S1 and S2 amplitudes due to their closer proximity to the atrioventricular valves and reduced attenuation through surrounding tissues. In contrast, recordings from the aortic and pulmonic areas generally show lower amplitudes because of greater distance from the primary sound sources and increased damping effects from intervening anatomical structures [54]. Thus, a randomly selected 15-second segment from the mitral and tricuspid areas was used for analysis to improve FHS segmentation performance.

### 2.3 Frequency Analysis of PCG Signals

Analogous to [54], the PCG data preprocessing steps included spike removal, resampling, segmentation, and normalization. The recordings were resampled to 2kHz, and a zero-phase Butterworth bandpass filter of [25, 800] Hz was applied [27, 38, 54, 55]. Then the FHS including S1, S2, systolic and diastolic phases, were extracted from segmented single heartbeats using the HMM-based method described in [52]. The frequency spectra were estimated using Welch’s power spectral density (PSD) estimate using a Hamming window of length 128ms, analogous to [53]. For each FHS segment, the dominant frequency was determined per subject. Candidate PSD peaks were validated against a Gaussian spectral profile with a minimum 10Hz bandwidth. This morphological constraint ensures the selection of physiologically relevant peaks, [53] while rejecting spurious, narrow-band spikes.

Mathematically, for an input PCG signal *x*[*t*] that has length *t*, let *s*[*n*] be a single heartbeat signal extracted from *x*[*t*]. And *s*[*n*] is divided into *M* valid overlapping frames of length *L* given by:

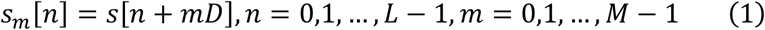

where, *s*_*m*_[*n*] is the *m* ^th^ segment, *D* is the step size between consecutive frames. The periodogram of each windowed segment, *P*_*m*_ (*f*), with a window function ω is given by:

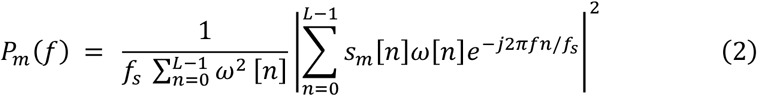

Then the Welsh PSD estimate, 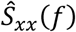, is obtained by averaging over all segments: as:

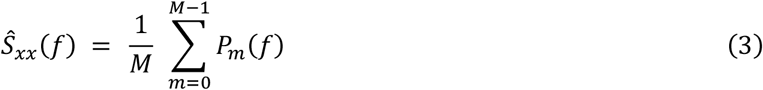

Further analysis on different bands based on the systolic and diastolic murmurs spectral ranges was also performed. Individual heartbeats consisting of all the FHS were extracted using [52], and frames were extracted with a 50% overlap. Four bands were defined, [25,100] Hz, [100,200] Hz, [200,400] Hz, and [400,800] Hz. The choice of these bands is based on the spectral content of heart sound components [53, 54, 55]. Note that, the fundamental frequency corresponding to each FHS segment estimated using 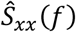, implicitly assumes the stationarity constraint of the segments, for instance systolic or diastolic segments.

However, the entire PCG waveform over long period is non-stationary [55]. A single global PSD estimate collapses the spectral temporal information into one average thereby losing the time-localized information associated with the FHS. Therefore, a band-limited time-varying spectral analysis based on the Short-Time Fourier Transform (STFT) was used. This enables to track spectral energy variations per the frequency bands [*f*_low_, *f*_high_], across the cardiac cycle. This can be computed by integrating the spectral energy across the corresponding frequency bins.

Let |*X*(*s*_*m*_, *k*)| denote the FFT coefficients of frame *s*_*m*_[*n*] and frequency bin *k* . The corresponding short-time spectral power (spectrogram) is defined as:

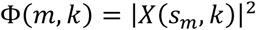

Hence, the spectral energy within each frequency bands [*f*_low_, *f*_high_], was computed over time by integrating the spectral power across the corresponding frequency bins:

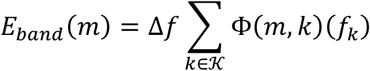

where *K* = { *k* | *f*_*low*_ ≤ *f*_*k*_ ≤ *f*_*high*_ } denotes the set of frequency-bin indices within the selected band, and 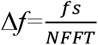 is the frequency resolution determined by the sampling frequency *f*_*s*_ and FFT length *N*_*FFT*_ . The quantity *E*_*banns*_ (*m*) therefore estimates the time-varying spectral energy within the selected frequency band at frame *s*_*m*_[*n*].

Statistical comparisons the relative band energies, *RE*_*bann*_, between the normal and PwRHD groups were performed for each frequency band. The relative spectral energy for each band was computed by normalizing the specific band energy, *E*_*b*_, with respect to the total spectral energy across all analyzed frequency bands:

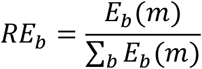

### 2.4. Classification Workflow

The classification workflow block diagram is provided in Figure 2. After filtering and preprocessing both the ECG and PCG signals, as explained in section 2.3, the features were extracted from a 15-second waveform segment of the input ECG or PCG signals. The list of features computed for PCG are provided in Table 1. The ECG features were computed analogous to our earlier work in [56]. The extracted features were evaluated for classification using XGBoost model.

**Table 1:** Extracted features from a PCG signal.

| Group | Feature | Reference |
| --- | --- | --- |
| Time-domain | Envelope | [26, 53] |
|  | Amplitude | [26, 53] |
|  | Energy | [26, 53] |
|  | Shannon Entropy | [33] |
|  | Autocorrelation | [26, 54] |
|  | Loudness | [29, 53] |
|  | Teager energy | [33] |
| Spectral | Dominant frequency (wave-form) | [27, 30] |
|  | Dominant frequencies (FHS) | [30] |
|  | Power variances | [34] |
| Mel-frequency | Log Mel Spectrum | [35] |
|  | MFCCs | [29, 35] |
| | $\Delta$, $\Delta^2$ | [29, 35, 68] |
| Wavelet | DWT | [30] |
| Non-linear | Bispectral statistics | [61] |
|  | Lyapunov exponent | [32, 69] |
|  | Fractal dimension | [62, 70] |
| Graph features | NVG | [62] |
|  | HVG | [62] |

**Figure 2.**
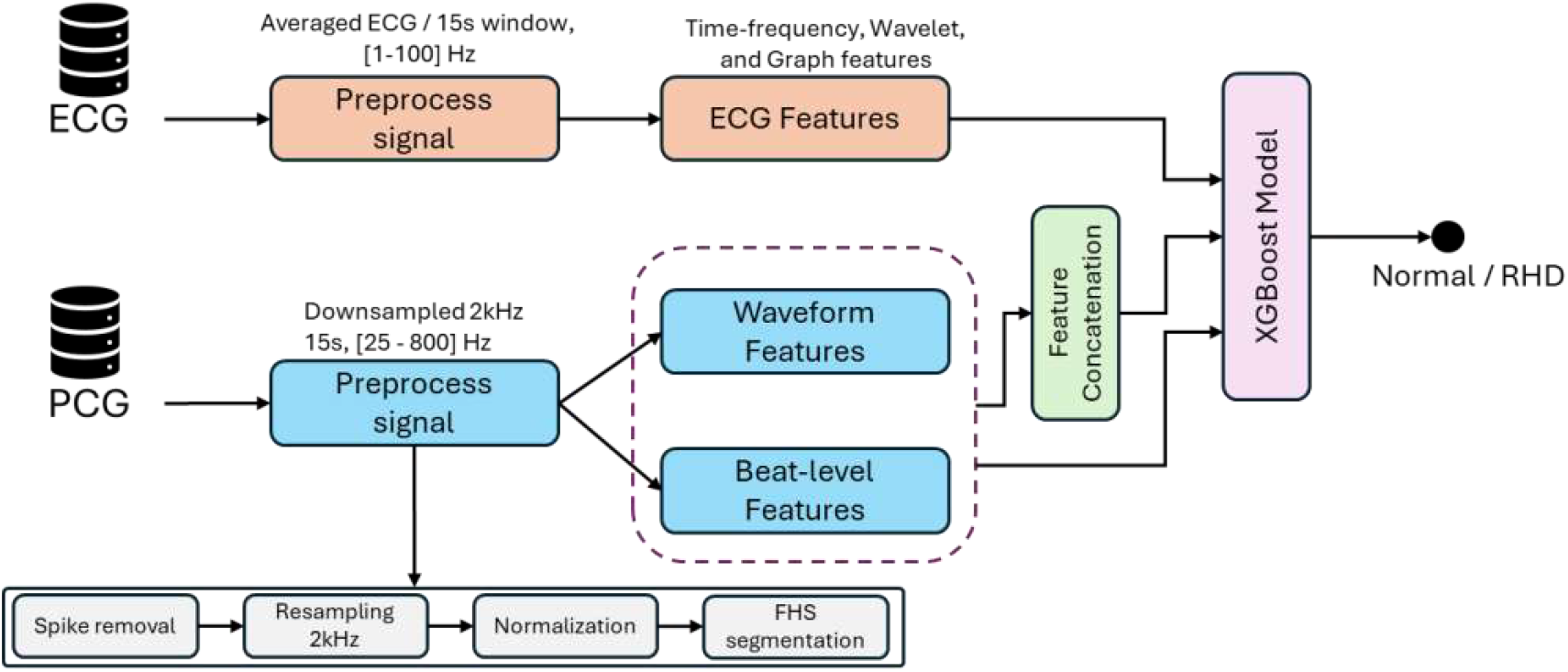
Workflow diagram of the classification task from multimodal ECG and PCG signals.

### 2.3 Feature Extraction

The extraction of meaningful features that capture the time-frequency and morphological patterns from the PCG signals were collected from previous works [50, 57, 58, 59]. The low-level features were derived from a short-time analysis of the signal and represent local, segment characteristics of fundamental sounds. These includes amplitude, envelope, autocorrelation, loudness, energy, and entropy. Moreover, statistical features that quantify global temporal behaviour through statistical moments such as mean, standard deviation, and skewness were also calculated.

The time-domain features are frequently utilized due to their capacity to accentuate temporal patterns and acoustic variations associated with physiological events, such as the FHS, murmurs, or external noises. Frequency-domain features, including Fourier-based spectral features and wavelet coefficients, provide insights into the signal’s periodicity and resonance properties.

The dominant spectral characteristics of the FHS across the bands described in Section 2.3, as well as the relative spectral power variances between S1 and systole, S2 and diastole, and systole and diastole, are also calculated.

Similar to [57, 58], the MFCCs along with their first order (*Δ*) and second-order (*Δ*2) derivatives computed using 13 Mel bands. Furthermore, similar to the work in [61, 62] non-linear features such as the Lyapunov exponent related to chaotic or dynamicity of the system using manifold decomposition was extracted. Visibility graph (VG) features from both natural VG (NVG) and horizontal VG (HVG) [63] were extracted for each band power graph from a single beat. These VG features include graph density, degree entropy, assortativity, and average path.

### 2.4 Model Evaluation Metric

In order to evaluate the discriminative power of the extracted features, we followed the methodology from our previous work [56]. The Extreme Gradient Boosting (XGBoost) classifier was employed, and the optimal hyperparameters were determined through a grid search over the parameter ranges described in Appendix A2. Hyperparameter optimization was performed using 5-fold stratified cross-validation with an F1-score as the optimization criterion.

The classification performance was evaluated using a 10-fold cross-validation. To ensure subject-level independence and prevent information leakage across splits, subject-stratified cross-validation was used. Considering the class imbalance in the dataset, class weights were computed from the sample sizes within each training fold. The ECG features from single-lead (lead II) and multilead (leads I, II, III, aVR, AVL, aVF) were fused with the PCG features. For multilead features, features from all leads were concatenated as one large input vector. In total 373 ECG features, 184 PCG features, and 3 metadata features were extracted per each subject.

Class weights were computed dynamically during each training fold based on class frequencies to mitigate the class imbalance. The positive predictive value 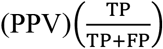, Sensitivity 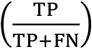, Specificity 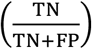, and 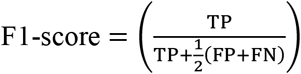 metrics were used to evaluate the model performance. Where TP is true positive, TN is true negative, FP is false positive, FN is false negative predictions of the model. In addition, to simulate the trade-offs between the available medical resources for confirmatory check and the target sensitivity to detect the RHD cases in the Global-South, class prediction probability score was thresholded between 50-80%. Furthermore, the impact of features that contributed to the model were evaluated using SHapley Additive exPlanations (SHAP) values [64].

## 3. Results

The aggregated dominant frequency for the FHS between Normal and PwRHD groups is shown in Figure 3. The frequency ranges for S1 and S2 are in the normal limits [54] for both groups. And no statistically significant mean differences was observed for systole (129.7 Hz vs. 129.8 Hz;t = 0.15, p= 0.88) and diastole (130.2 Hz vs. 131.1 Hz; t = 0.92, p = 0.36) phases. In addition, comparison of the PSD estimate of frames from individual beats over time resulted in slightly varied relative temporal energies of the band powers, *RE*_*b*_, between the two groups, particularly at the band [25-100] Hz.

**Figure 3.**
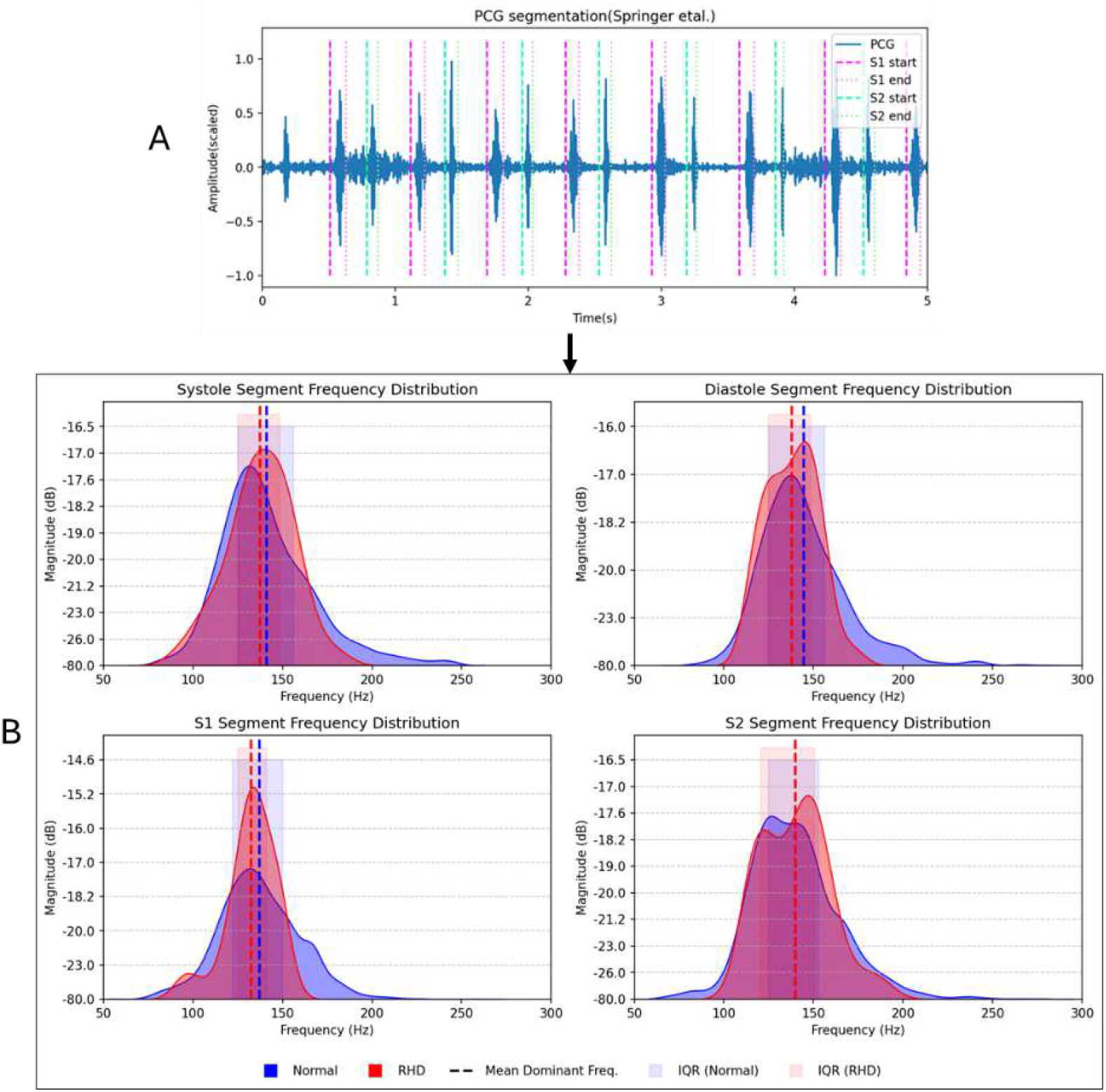
Extraction of FHS and its dominant frequencies: in panel (A) segmentation of FHS using the Springer’s algorithm [52] for a 5-second example signal. In panel (B) kernel density distributions of dominant frequencies for each FHS segments (S1, S2, systole, diastole) in Normal and RHD classes in asymptomatic cohort. Vertical dashed lines denote mean frequencies per class, and shaded regions represent ±1 standard deviation around the mean.

However, the mean relative energy in the high-frequency bands ([100–200] Hz and [200– 400] Hz), of the S1 segments did not differ significantly between the two groups, with values of 46.9% vs. 47.0% (p = 0.995) and 46.1% vs. 46.8% (p = 0.445), respectively. Similarly, no significant differences were observed for the S2 segments, with relative energies of 23.2% vs. 21.9% (p = 0.104) in the [100–200] Hz band and 26.9% vs. 27.3% (p = 0.725) in the [200– 400] Hz band. The remaining relative energy was concentrated in the [25–100] Hz band, and differed significantly only for the S1 segments (29.6% vs. 31.2%; p = 0.021).

The classification performance of the XGBoost model for the features from ECG, PCG, and combined ECG and PCG datasets is shown in Table 2. The model evaluation on ECG features alone yielded the best results with a sensitivity of 61.6%, a specificity of 97.3%, a PPV of 64.1%, and an F1-score of 61.1%. In contrast, features from PCG alone showed poor performance, with a sensitivity of 46.8%, a specificity of 68.8%, a PPV of 11.1% and an F1-score of 23.5%. As shown in Table 2, the combined ECG and PCG features yielded a sensitivity of 59.6%, a PPV of 63.6%, and an F1-score of 60.8%. The PCG combined with multilead ECG features showed relatively better results than the ECG features combined with single-lead (Lead II) ECG. In addition, the AUROC in Figure illustrates the behaviour of each modality and their fusion where a nearly random performance, (AUROC=51.3), was observed from PCG features.

**Table 2:**
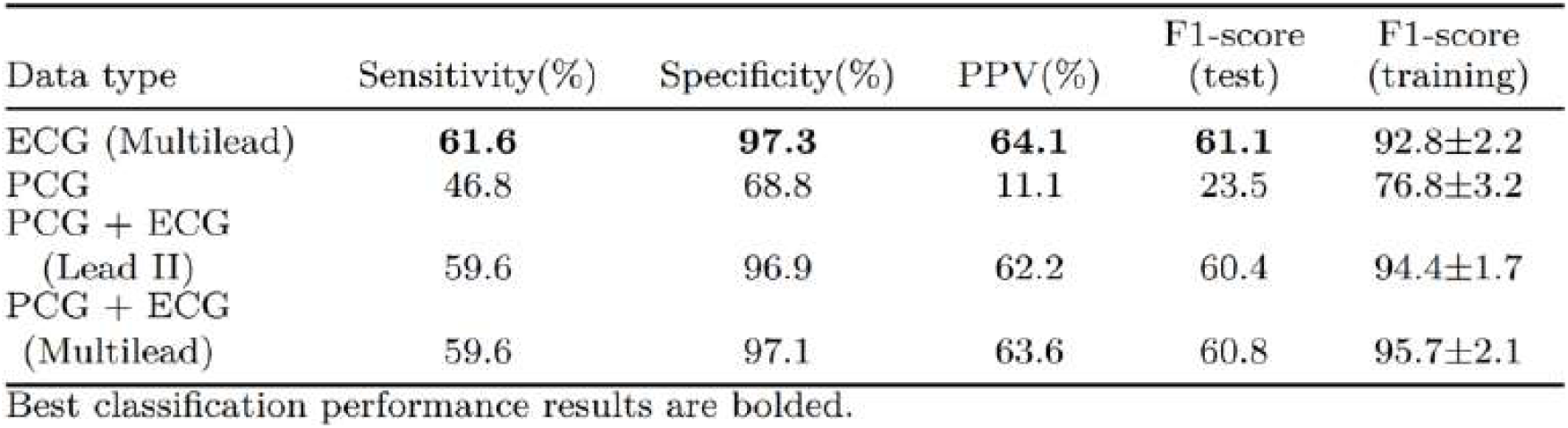
10-folds classification results on ECG, PCG, and combined features of both ECG and PCG.

Examining the model’s performance at various sensitivity values by changing class probability score thresholds revealed that combining multimodal features did not improve performance over ECG alone. The precision-recall (PR) curve illustrated in Figure 4 shows the 10-fold results of the model at different class probability threshold values. The performance was poor at higher sensitivity thresholds above 70% sensitivity. An F1-score of 35.9% at 70% sensitivity, 27.2% at 80% sensitivity, and 18.1% at a sensitivity of 90%. However, mediocre performance were observed at a sensitivity range between 50-60%, with an F1-score of 60.8% at 59.6% sensitivity.

**Figure 4.**
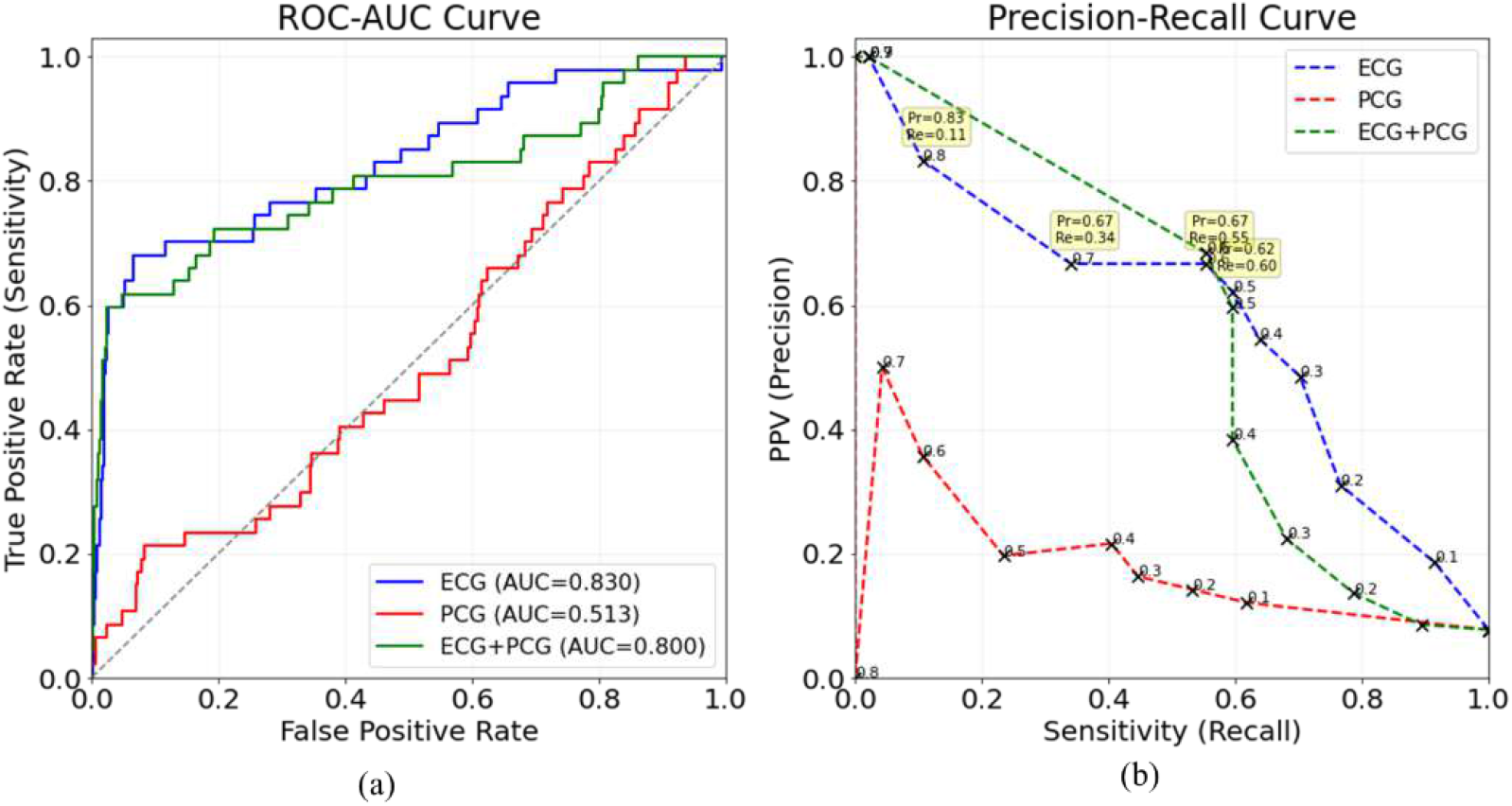
(a) The ROC curves and (b) Precision–recall curves of the XGBoost models trained using ECG features, PCG features, and combined ECG and PCG features.

Feature contributions calculated from SHAP values is shown in Figure 5. The list of features that impacted the model prediction most are mixed from both feature modalities, but majority are from ECG features than PCG features. From PCG the graph-features, and MFCCs were predominantly impactful. Whereas from ECG time domain features from the QRS and T-wave, and from the graph features were the most impactful. In addition, from the meta features, Sex also contributed to the model’s decision.

**Figure 5.**
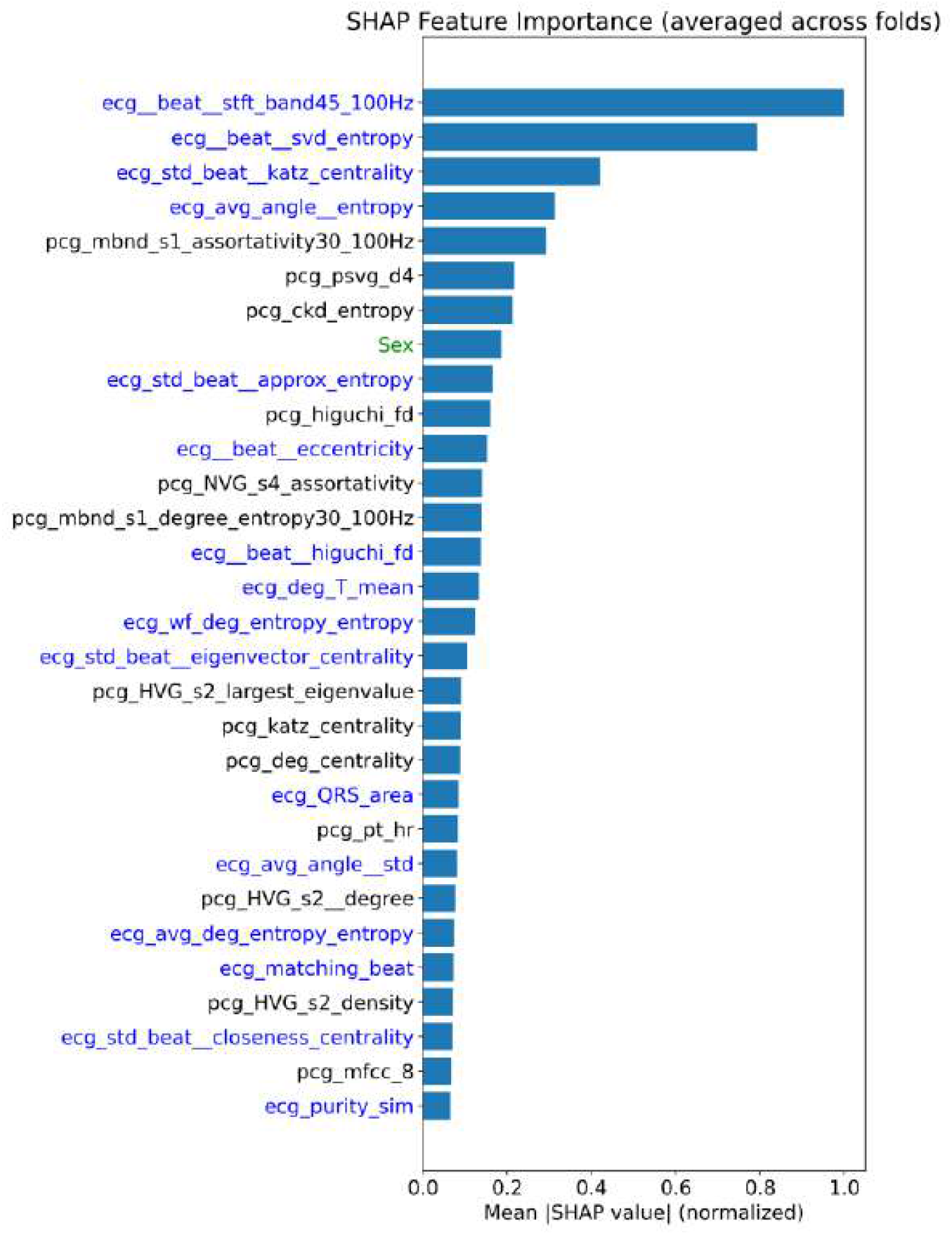
SHAP summary plot for combined features. The colors encode the feature domains (blue: ECG features, black: PCG features, and green: metadata). The x-axis normalized mean feature importance, and y-axis shows the contributions of the top 30 features. Explanation of these features were given in Appendix A3.

## 4. Discussion

This study evaluated the effectiveness of combined features extracted from ECG and PCG signals for an early detection of RHD among asymptomatic schoolchildren. The findings demonstrated that the multimodal approach did not show better results than those achieved using ECG alone.

### 4.1. Performance Comparison of Modalities

As described in Table 2, the model performance based on PCG features was poor, achieving an F1-score of 23.5% and a sensitivity of 46.8%. The asymptomaticity of the study cohort which resulted in faint murmurs for PwRHD, class imbalance of the dataset, and inherent noises in the PCG recordings were the main factors for the obtained results. The observed gap between training and test F1-scores (see in Table 2) indicates a bias–variance trade-off, despite the use of tuned best hyperparameters and class weights. In particular, the model achieves consistently high training performance but substantially lower test performance, suggesting high variance. This is primarily attributed to the extreme class imbalance between healthy participants and PwRHD, which limits the model’s ability to learn robust minority-class decision boundaries. In such screening settings, generalization to unseen data is degraded.

Nevertheless, our findings are in line with the findings from [65], where early rheumatic valvulitis may not produce loud murmurs. Such early RHD associated murmurs are often soft, medium intensity, and inconsistent [65]. Clinically, the murmur intensity correlates with severity of the underlying regurgitation or stenosis [1,54]. Consequently, as described in Figure 3, the mean fundamental frequency of the FHS for systolic and diastolic phases are not statistically significantly differed for the Normal and PwRHD groups.

Similar low detection performance results using PCG were also reported by [9, 11], showing that the use of auscultation for detecting subclinical or early-stage RHD lesions showed low sensitivity <50% when valuated on echocardiography-confirmed cases. Furthermore, the study in [11, 12] showed the sensitivity of auscultation alone to detect pathological murmurs was 4-46.4%, missing silent but pathologic valvular abnormalities in both definite RHD and borderline RHD. Auscultation-based RHD screening performance among children has been reported to be low, with a sensitivity of 38.2%, a specificity of 75.1%, and a PPV of 5.1% [13].

In contrast, the ECG-based features showed significantly better results compared to PCG-based results. As shown in Table 2, the ECG features alone achieved a sensitivity of 61.6%, a specificity of 97.3%, and a PPV of 64.1%. Since early RHD involves myocarditis and conduction system inflammation before structural valve disease becomes hemodynamically significant [1], evidence of conduction involvement early in the disease was also indicated [11, 13, 57]. The ECG captures the electrical consequences of such inflammation, hence the better result than pronounced mechanical problems captured using the PCG. Moreover, from [24, 38] the relatively better SNR and temporal conduction variations in ECG wave morphologies contributed as well.

However, there were relatively similar results observed from multimodal features when compared with the ECG-only features on overall classification. The inclusion of PCG modality did not show improved results, as shown in Table 2. This poor discriminative performance suggests that the insignificant differences in spectral contents of the FHS, and hence weak acoustic characteristics of murmurs in early asymptomatic RHD. Although the PCG features were among the top important features (see Figure 5), they showed no performance gains over the ECG alone.

#### 6.4.2 Clinical implications

The global epidemiological reports consistently emphasize that early disease detection at large scale is a primary barrier to control RHD rather than treatment availability [3]. In RHD endemic high prevalent regions, moderate-to-high sensitivity is prioritized over perfect specificity since the screening objective is early identification rather than definitive diagnosis. However, in regions having constrained medical resource setting, the sensitivity (see Figure 4(b)) can be adjusted to moderate levels without overwhelming the confirmatory procedures. As indicated in our previous work [56], such moderate performance was observed at threshold of 0.6%.

From a modality point-of-view, the ECG is better suited for scalable and ease of use deployment. It requires no expertise for acquisition with a non-intrusive manner hence aligns with the efforts on shifting the population screening task to non-professional local personnel. In contrast, phonocardiography relies on acoustic manifestations the valve closures hence the recording sensor needs to be positioned at precise locations, and require understanding of anatomical knowledge with careful handling of the stethoscope. Thus, a trained healthcare workers are necessary for PCG acquisition.

## 5. Limitations

Nonetheless, this study has several limitations. First, the small sample size may lead to overfitting and reduced generalizability, particularly across diverse populations and recording environments. Second, a lack of large-scale, standardized public datasets hinders external validation and the benchmarking of deep learning models. Finally, while our framework utilizes advanced time–frequency and visibility-graph features, these complex representations lack clinical interpretability. Future research should prioritize larger, multimodal datasets and foundation models to improve both robustness and model explainability for the clinicians.

## 6. Conclusion

This study evaluated a comprehensive set of features from ECG and PCG signals to investigate the potential of low-cost alternative devices for screening asymptomatic RHD using machine learning. The performance of PCG alone for the automated early detection of asymptomatic RHD among at-risk schoolchildren is poor, with an F1-score of 23.5%, compared to ECG, which achieved 61.1%. In contrast to the findings in advanced RHD, the spectral content of systolic and diastolic segments showed no statistically significant differences between the healthy and RHD groups. Consequently, adding PCG to ECG does not improve model performance, as the PCG contributes less-distinctive features that fail to enhance class separability. Early disease causes only subtle or faint murmurs that overlap with normal heart sounds, making it hard to extract useful features across schoolchildren and adolescents.

In settings where resources are limited and RHD is endemic, ECG with automated machine learning can be a more effective, low-cost screening method for asymptomatic RHD than phonocardiography. These findings support WHO-aligned RHD strategies that prioritize early detection through decentralized, school-based screening as an initial triage step. Individuals identified as high risk for RHD by the model can subsequently be referred for confirmatory echocardiography. Such low-cost screening is suitable for task-shifting, where trained non-physician personnel can conduct screening with the support from automated machine learning–based interpretation.

## Data Availability

The dataset used in this study is not publicly available, but can be accessed with a reasonable request to the corresponding author.

## 7. Declarations

### 7.1 Ethics approval and consent to participate

The study was conducted according to the guidelines of the Declaration of Helsinki and approved by the Ethics Committees of the UZ Leuven with reference No: B3222022001075, Soddo Christian Hospital No:SCH1941015. Clinical trial number is not applicable.

### 7.2 Consent for publication

Written informed consent was obtained from all the patient(s) to publish the study results.

### 7.3 Availability of data and materials

The RHD dataset analyzed in this study is not publicly available. However, it can be shared with a reasonable request via https://doi.org/10.48804/HIROCQJ.

## 7.6 Acknowledgment

The authors are grateful to the RHDECG dataset collaborators, the study participants, the schools and hospitals involved, and the examining physicians during the study campaign. We also extend our gratitude to the Flanders AI program for supporting this study.

## Abbreviations

RHD: Rheumatic heart disease
ARF: Acute Rheumatic Fever
ECG: Electrocardiogram
PCG: Phonocardiogram
Pw: P-wave
Tw: T-wave
QRSw: QRS complex (QRS wave)
DL: Deep Learning
ML: Machine Learning
KM6L: Kardia Mobile 6L ECG sensor
DNN: Deep Neural Network
CNN: Convolutional Neural Networks
LSTM: Long Short-Term Memory

## Appendix

**Table A1:**
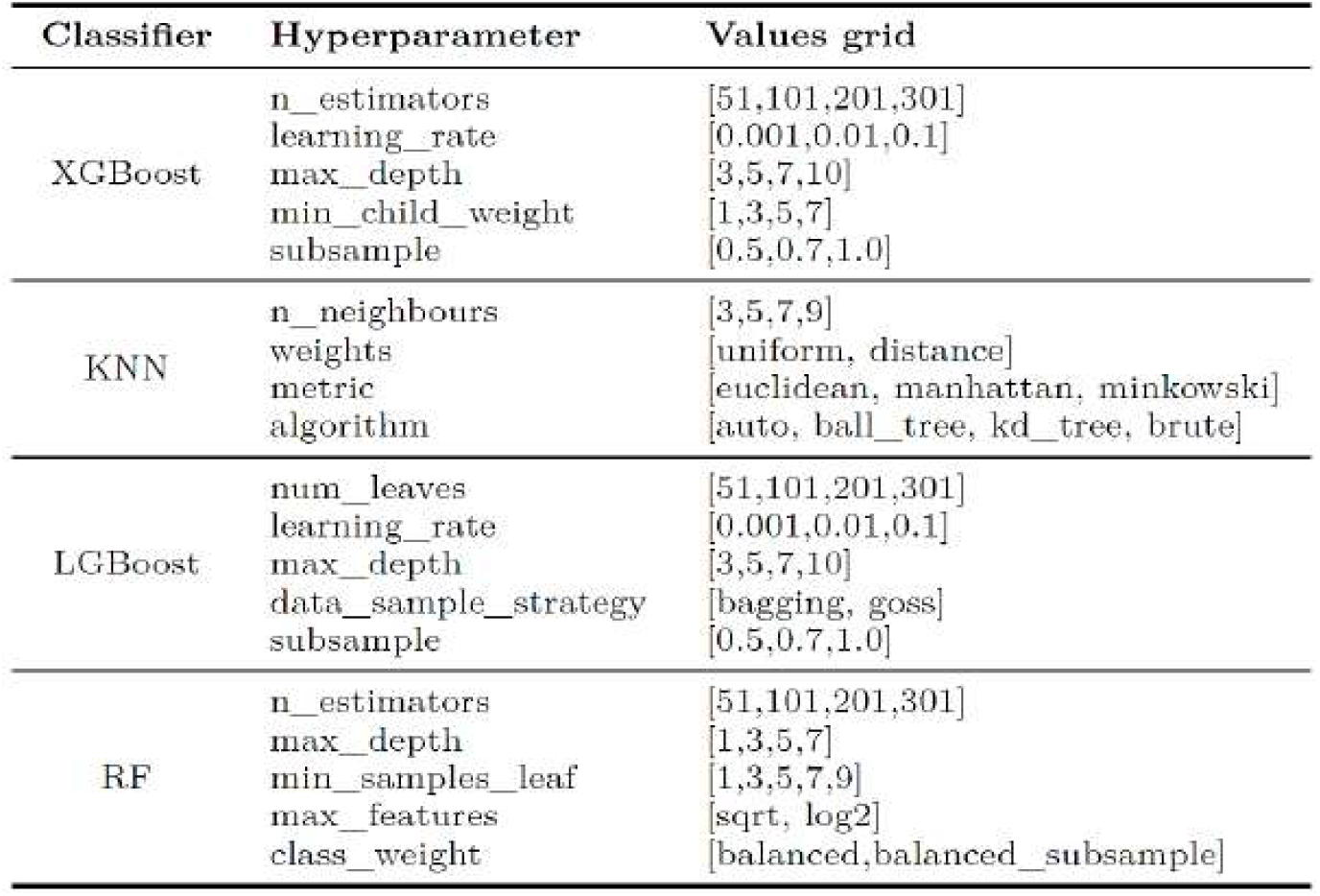
Hyperparameters grid table for different ML classifiers.

### A2: SHAPely Feature Importance

The SHAP computes how much each of these features individually impacts the model output toward predicting “Normal” or “PwRHD” classes, while accounting for interactions with other features by evaluating various feature combinations. Mathematically, for a given *i*^th^ feature vector row of *F*_*i*_ *=* {*v*_*1*_, *v*_*2*_, …, *v*_*m*_} with its class prediction *f*_*θ*_*(F*_*i*_), SHAP calculates a marginal contribution of each feature *v*_*i*_ to the model output by comparing what the model would predict with and without that feature, averaged across all possible feature combinations. Thus, the SHAP value *ϕ*_*i*_ for a feature *v*_*i*_ is calculated as:

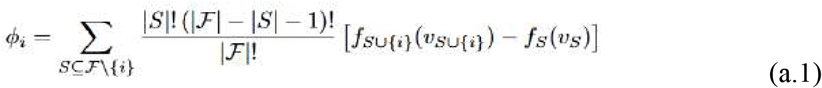

where ℱ is the set of all input features, *S* is a subset of features not containing *v*_*i*_, *f*_*S*_ *(v*_*S*_ ) is the model’s prediction using only features in subset *S, f*_*S*_∪{*i*}(*v*_*S*_∪*{i}*) is the prediction when feature *v*_*i*_ is added to subset *S*. In order to get global importance of each *i*^*th*^ feature, SHAP values can be aggregated across the row feature samples, *R*, which is given as:

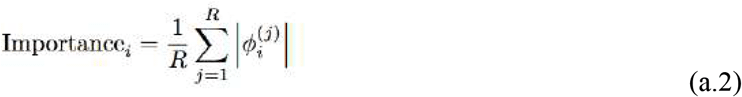

Since *ϕ*_*i*_ indicates how much feature *v*_*i*_ contributed to the prediction for that specific sample, to get rid of the direction of influence its magnitude is considered. So, equation (a.1) computes local SHAP contribution of a feature for one prediction. Whereas equation aggregates those local contributions into global feature importance. Hence, the importance of the classifier illustrated in Figure 5 is computed.

**Table A2:** Performance comparison of different machine learning models on multimodal ECG and PCG features.

| Model | Sensitivity(%) | Specificity(%) | PPV(%) | F1-score(%) |
| --- | --- | --- | --- | --- |
| XGBoost | 59.6 | **97.1** | **63.6** | **60.8** |
| LGBoost | **72.3** | 57.9 | 12.5 | 43.6 |
| Random Forest | 59.6 | 95.9 | 54.9 | 58.8 |
| KNN | 44.6 | 70.0 | 11.1 | 31.4 |

The best results were in bold.

Table A4 shows the performance of combined features evaluated using different classifiers. The optimal hyperparameters were tuned similar fashion with the XGBoost (see Section 2.4 from grid of hyperparameter values listed in Table A1 for k-nearest neighbours (KNN), Light Gradient Boosting (LGBoost), and Random Forest (RF) classifiers. Consistent performance trends were observed with different ML classifiers. Table A2 shows the performance of different ML classifiers on the multimodal features. The KNN showed the lowest performance with an F1-score of 31.1%, a sensitivity of 44.6% and very low PPV of 11.1%. The RF models showed comparable results with the XGBoost, achieving an F1-score of 58.8% with a sensitivity of 59.6%. Whereas, the LGBoost has slightly lower results across all metrics, but a high sensitivity of 72.3%. Overall, the XGBoost yielded best results than these all the other classifiers.

#### A3: Definitions of the Features

- Time-dimain:
  - *ecg_beat_svd_entropy*: average entropy of Singular Value Decomposition (SVD) of 1-second window.
  - *ecg_std_beat_approx_entropy*: Standard deviation of entropy across beats. Measures variability in beat unpredictability.
  - *ecg_QRS_area*: area under the QRS complex of an averaged ECG beat.
  - *ecg_matching_beat*: autocorrelation between current beat and average beat.
  - *pcg_ckd_entropy*: log-amplitude complexity measure of the PCG signal.
  - *pcg_pt_hr*: average heart rate estimated from PCG envelope.
- Frequency-domain
  - *ecg_beat_stft_band45_100Hz*: The spectral energy contained in the [45–100]Hz frequency band of the Short-Time Fourier Transform (STFT) of an ECG beat. Measures the distribution of signal energy in the high-frequency range of the ECG beat over time.
  - pcg_mfcc_8: the 8th MFCC of the PCG signal.
- Non-linear
  - *ecg_beat_higuchi_fd*: Fractal dimension of ECG waveform using Higuchi’s method. Measures waveform complexity across scales.
  - *pcg_higuchi_fd*: the Higuchi fractal dimension of the PCG signal. Measures waveform complexity across scales.
- Visibility Graph
  - *ecg_std_beat_katz_centrality*: Standard deviation of Katz centrality values extracted from Natural Visibility Graph (NVG) across beats. Measures variability in graph connectivity structure.
  - *ecg_avg_angle_entropy*: Average entropy of edge angles in the NVG. Measures randomness in directional structure/slopes of the ECG graph.
  - *ecg_beat_eccentricity*: Graph eccentricity metric computed on NVG of a beat. eccentricity(node) = max shortest-path distance to other nodes. Captures how spread/elongated the graph structure is.
  - *ecg_beat_deg_T_mean*: average node degree over T-wave segments. Measures average graph connectivity density across T-waves.
  - *ecg_beat_wf_deg_entropy*: Entropy of node degree distribution in NVG windows. Measures randomness of connectivity patterns.
  - *ecg_std_beat_eigenvector_centrality*: Standard deviation of eigenvector centrality across beats. Measures variability of dominant graph-node influence patterns.
  - *ecg_avg_angle_std*: standard deviation of average graph angles.
  - *ecg_avg_deg_entropy*: average entropy of graph degree distributions of the ECG signal. Measures complexity/randomness of connectivity.
  - *ecg_std_closeness_centrality*: standard deviation of closeness centrality values from NVG of the ECG signal. Measures how near nodes are to all others to capture variability in graph connectivity.
  - *pcg_mbnd_s1_assortativity30_100Hz*: assortativity coefficient computed from the NVG of the S1 heart sound segments in the [30–100] Hz frequency band. Measures structural organization of S1 dynamics in the 30–100 Hz band.
  - *pcg_psvg_d4*: the fourth power of scale-freeness in visibility graph (PSVG) diagonals.
  - *pcg_NVG_s4_assortativity*: assortativity coefficient of the NVG constructed from diastolic segments.
  - *pcg_mbnd_s1_degree-entropy30_100Hz*: Entropy of the node degree distribution of the NVG from the S1 segments in the [30–100] Hz band.
  - *pcg_HVG_s2_largest_eigenvalue*: the largest eigenvalue of the adjacency matrix of the Horizontal Visibility Graph (HVG) for S2 segments.
  - *pcg_katz_centrality*: Katz centrality computed on the PCG visibility graph.
  - *pcg_deg_centrality*: average Degree centrality of nodes in the PCG graph.
  - *pcg_HVG_s2_degree*: Average or total node degree in the HVG for segment s2.
  - *pcg_HVG_s2_density*: Graph density of the HVG for segment s2.

